# Speech Acoustic Markers Detect APOE-ε4 Carrier Status in Cognitively Healthy Individuals

**DOI:** 10.1101/2025.10.03.25337276

**Authors:** Maryam Tavakoli, Mehrdad Dadgostar, Jordan R. Green, David H. Salat, Steven E. Arnold, Kathryn P. Connaghan, Brian Richburg, Nelson Barnett, Mariam Tkeshelashvili, Marziye Eshghi

**Affiliations:** MGH Institute of Health Professions, Charlestown, MA, USA; Athinoula A. Martinos Center for Biomedical Imaging, Harvard-MIT Health Sciences and Technology, Charlestown, MA, USA; Speech and Hearing Bioscience and Technology, Harvard University, Boston, MA, USA; Massachusetts General Hospital, Department of Radiology, Harvard Medical School, Boston, MA, USA; Neuroimaging Research for Veterans Center, VA Boston Healthcare System, Jamaica Plain, MA, USA; Massachusetts General Hospital, Department of Neurology, Harvard Medical School, Boston, MA, USA

**Keywords:** APOE-ε4 genotype, genetic risk factor, Alzheimer’s disease, Speech biomarkers, Speech Acoustics, eGeMAPS

## Abstract

**Background:** APOE-ε4, the strongest genetic risk factor for Alzheimer’s disease (AD), is linked to early motor vulnerability, including subtle speech control changes. Because speech integrates fine neuromotor processes, acoustic analysis could offer a sensitive, noninvasive marker of preclinical effects.

**Objectives:** To determine if speech acoustics distinguish cognitively healthy APOE-ε4 carriers from non-carriers, and to assess which speech tasks provide optimal classification performance.

**Design:** A cross-sectional observational study employing supervised machine learning to analyze acoustic features extracted from multiple speech tasks. Genetic algorithms (GAs) were used for feature selection, and model performance was compared across task contexts.

**Setting:** All assessments and recordings were conducted in a sound-attenuated laboratory at MGH Institute of Health Professions.

**Participants:** Forty-four cognitively healthy adults (19 APOE-ε4 carriers, 25 non-carriers), aged 57–79 with no history of neurological and psychiatric conditions.

**Measurements:** Digitized speech was analyzed for 88 eGeMAPS acoustic features. Random Forest classifiers were trained to distinguish genotypes; model optimization employed GAs and stratified cross-validation. Performance was evaluated using F1 scores and subgroup analyses for sex effects.

**Results:** Random Forest classification of spontaneous speech achieved F1 scores above 0.90 for distinguishing APOE-ε4 carrier status, outperforming performance on structured tasks. GA-based feature selection consistently improved classification. Accuracy was highest among female participants. The combined speech dataset confirmed the robustness and generalizability of results.

**Conclusions:** Automated analysis of speech acoustics especially from spontaneous speech, detects APOE-ε4 carrier status in asymptomatic adults, supporting speech as a scalable digital biomarker for early Alzheimer’s risk.

## Introduction

Alzheimer’s disease (AD), the leading cause of dementia worldwide, has a prolonged preclinical phase during which pathophysiological changes accumulate in the absence of cognitive symptoms [1]. Because neurodegeneration begins years before symptoms emerge, delaying intervention until clinical decline is apparent sharply limits the effectiveness of available treatments and trials. Detecting individuals at this asymptomatic stage remains a major clinical challenge, underscoring the need for scalable, non-invasive biomarkers that can identify at-risk individuals prior to symptomatic onset. Current biomarker approaches such as cerebrospinal fluid analysis and PET imaging, while informative, are invasive, costly, and not scalable for routine screening. Therefore, it is necessary to identify and validate alternative approaches that are accessible, low-burden, and deployable in large-scale or remote settings.

The apolipoprotein E ε4 (APOE-ε4) allele is the strongest genetic risk factor for late-onset AD [2] and has been linked to an accelerated rate of cognitive and motor decline in aging populations[3–5]. While the association between APOE-ε4 and memory decline is well established [6, 7], emerging evidence suggests broader effects on cognition, including impairments in language function and executive control, in older adults [8–10]. Longitudinal studies indicate that ε4 carriers experience more rapid deterioration in verbal fluency and complex cognitive tasks compared to non-carriers [10]. In parallel, motor dysfunction associated with APOE-ε4 has been observed in both gross and fine motor control, including speech motor function [4, 11–13].

Speech and language changes are increasingly recognized as early markers of AD pathology [14–16]. As speech integrates motor planning, executive control, language processing, and memory, it provides a multidomain indicator of brain health and an ecologically valid diagnostic window. Individuals with mild cognitive impairment (MCI) and AD frequently exhibit slowed speech tempo, increased pausing, and reduced semantic complexity [15, 17–19]. Recent machine learning studies have leveraged these features to discriminate MCI and AD, achieving accuracies up to 88% in diverse cohorts [20, 21]. However, most prior research has focused on language– and cognition-based markers. Because these markers reflect downstream cognitive dysfunction, they often detect pathology only after cognitive symptoms are visible. In contrast, speech features reflecting motor control (e.g., prosody and tempo) may offer a more sensitive window into early, preclinical disease processes, as neuromotor changes can precede overt cognitive decline. Nonetheless, research specifically targeting motor-based speech changes in cognitively healthy but genetically at-risk groups, such as APOE-ε4 carriers, remains very limited.

Emerging evidence shows that motor function declines more rapidly in individuals with the APOE-ε4 allele than in non-carriers [4, 22]. This includes losses in muscle strength, gait stability, and coordination, making ε4 carriers an important group for studying neuromotor vulnerability before cognitive symptoms arise. Speech provides a particularly sensitive window into these changes as ε4 carriers exhibit abnormal orofacial muscle recruitment during speech, with increased amplitude, frequency, and synchrony of motor unit activation [11]. These neuromuscular differences can predict ε4 carrier status with up to AUC = 0.90, outperforming cognitive tests [11]. In addition, speech kinematic features including altered lip movement duration, speed, and range can differentiate cognitively normal ε4 carriers from non-carriers, capturing subtle abnormalities in motor control that may signal elevated AD risk [13]. Machine learning models trained on these kinematic features achieved up to 87.5% accuracy for APOE-ε4 classification, further supporting the utility of speech as a high-yield early biomarker [13].

Because speech sounds are produced through precise movements of the lips, tongue, and jaw, even small disruptions in neuromotor control are expected to propagate to measurable changes in acoustic features. While kinematic measures capture details of articulatory movement like the range and timing of lip and jaw motions, acoustic features reflect how these movements are realized in the speech signal, including qualities such as voice and timing. Importantly, kinematic data acquisition typically requires specialized equipment, controlled environments, and technical expertise for data capture and analysis, limiting its feasibility for broad implementation. In contrast, acoustic data can be collected easily and non-invasively using mobile or telehealth platforms, making it highly suitable for large-scale and at-home monitoring.

Advances in automated acoustic analysis and machine learning now enable objective and scalable evaluation of speech patterns, opening new avenues for remote risk screening in preclinical populations [23]. This study has two primary objectives: (1) to develop and validate an automated speech analytics pipeline that classifies cognitively healthy APOE-ε4 carriers and non-carriers using acoustic features from the extended Geneva Minimalistic Acoustic Parameter Set (eGeMAPS) [24], and (2) to identify the speech tasks that maximize classification accuracy and hence would be the most efficient task to be incorporated in clinical screening and remote data collection.

Different speech tasks elicit distinct acoustic-linguistic profiles due to their unique cognitive and motor demands; indiscriminate pooling of recordings across tasks can introduce heterogeneity that may obscure meaningful risk-related patterns. Thus, evaluating the ability of acoustic features to distinguish APOE-ε4 carriers across various speech tasks is essential for identifying protocols that are both diagnostically sensitive and logistically practical. Strategic selection and optimization of speech tasks is likely to support more accurate early risk detection than a comprehensive “all-in” approach, ensuring that assessments capture the informative nuances revealed by task demands.

To our knowledge, this is among the first studies to classify cognitively healthy APOE-ε4 carriers using eGeMAPS-derived features, highlighting their potential as non-invasive biomarkers of elevated AD risk before symptom onset. By systematically evaluating speech tasks, the study also provides guidance for developing efficient, accurate protocols tailored to both clinical and remote applications.

## Methods

The study was approved by the Mass General Brigham (MGB) Institutional Review Board (IRB Protocol #2021P001460). All procedures were conducted in accordance with MGB IRB guidelines and regulations. Written informed consent was obtained from all participants prior to their participation.

### 2.1 Participant population

A total of 44 participants (19 females, 25 males; age range: 57–79 years; mean age: 66.9 years) were enrolled in this study. Participants were divided into two groups based on their APOE genotype: APOE*-ε4* positive (ε4+) (n=19, 8 males, 11 females) and APOE*-ε4* negative (ε4-) (n=25, 17 males, 8 females). The ε4+ group included individuals with ε3/ε4 heterozygous and ε4/ε4 homozygous genotypes, while the ε4-group consisted of individuals with the ε3/ε3 homozygous genotype. The groups were matched for education (>12 years) and age (p-value=0.13), with age control being particularly important to minimize potential aging effects on speech outcomes. APOE genotyping was conducted using blood samples, and all participants were blind to their respective genotypes. Demographic and clinical information for both groups are summarized in Table 1.

**Table 1.** Demographic and clinical Characteristics Stratified by Genotype Carrier Status.

| Variables | All participants | $\epsilon 4^-$ | $\epsilon 4^+$ | *p*-value |
| --- | --- | --- | --- | --- |
|  | (*n* = 44) | (*n* = 25) | (*n* = 19) |  |
| Female, *n* (%) | 19 (43.18) | 8 (32.00) | 11 (57.89) | 0.085 <sup>†</sup> |
| Family history of AD, *n* (%) | 24 (54.55) | 14 (56.00) | 10 (52.63) | 0.824 <sup>†</sup> |
| Age, mean (*SD*) | 66.95 (5.80) | 68.08 (6.15) | 65.47 (5.09) | 0.132 <sup>‡</sup> |
| Total years of education, mean (*SD*) | 16.43 (2.10) | 16.84 (1.93) | 15.89 (2.23) | 0.149 <sup>‡</sup> |
| MMSE, mean (*SD*) | 29.11 (1.17) | 29.00 (1.32) | 29.26 (0.93) | 0.443 <sup>‡</sup> |
| MoCA, mean (*SD*) | 27.36 (2.26) | 27.40 (2.36) | 27.32 (2.19) | 0.903 <sup>‡</sup> |

MMSE = mini-mental state examination; MoCA = Montreal cognitive assessment; SD = standard deviation. <sup>†</sup> Chi-square test. <sup>‡</sup> t-test.

All participants met the following inclusion criteria: (1) capacity to provide informed consent and comply with task-specific study instructions; (2) native speakers of American English; (3) no previous record of speech, language, hearing, psychiatric, or neurological disorders; (4) no clinical diagnosis of MCI; (5) not currently using psychoactive drugs that could potentially impact study outcomes; (6) passed an age-adjusted pure-tone hearing screening, ensuring normal hearing acuity in at least one ear; and (7) dental and occlusion status within functional limits, as determined by an oral examination performed by ME. Additionally, all participants demonstrated normal functional speech as assessed through the Sentence Intelligibility Test (SIT), a standardized tool that measures speech intelligibility and speaking rate [25]. During the assessment, participants read 11 sentences randomly generated by the SIT software, with sentence length progressively increasing from 5 to 15 words. Their speech was digitally recorded and subsequently transcribed orthographically by two native American English-speaking listeners with established high interrater reliability (ICC= 0.99, p<0.001). The SIT software calculated speech intelligibility as the percentage of correctly transcribed words and speaking rate as the number of words read per minute (WPM). Furthermore, participants exhibited no sound distortions or abnormalities in voice, resonance, prosody, or rate qualities, as independently judged by two expert raters (ME and K P. C).

### 2.2 Experimental tasks

Participants were asked to produce speech stimuli varying in length and motor-cognitive complexity, including a sustained phonation task, syllable repetitions in an oral diadochokinetic (DDK) paradigm, passage reading, a picture description task, and spontaneous speech. In total, 340 voice samples were collected across all tasks, with 128 derived specifically from the spontaneous speech tasks. Each task is described in detail in Appendix 1.

### 2.3 Audio recording

All speech recordings were obtained in a sound-attenuated, acoustically treated room within the Speech Physiology and Neurobiology of Aging and Dementia (SPaN-AD) Laboratory to minimize ambient noise and reverberation. Participants were seated upright in a comfortable position, with a fixed head orientation and consistent distance from the microphone. Speech was captured using a high-fidelity, head-mounted condenser microphone (Williams AV MIC 094 Unidirectional Headband), positioned 10–15 cm from the mouth to ensure uniform signal quality. The microphone was routed through a Focusrite Scarlett 2i2 audio interface, with gain levels standardized and calibrated before each session. Recordings were digitized at 22.5 kHz with 32-bit resolution in uncompressed mono WAV format to maximize acoustic fidelity.

Prior to recording, participants were instructed to use their natural speaking voice at a comfortable loudness and pace while avoiding extraneous sounds (e.g., coughing, shuffling). Standardized seating, microphone positioning, and a preliminary calibration utterance were employed across participants to verify uniform input gain and consistent recording conditions. Quality assurance procedures included routine system calibration, real-time monitoring, and post-hoc inspection of waveforms and spectrograms for noise or artifacts. Any segments compromised by excessive noise or recording errors were re-collected to ensure data integrity.

### 2.4 Acoustic feature extraction

Digitized audio samples were post-processed in Audacity (Version 3.7.1, Audacity Team) to remove extraneous silences and non-speech segments. Each recording was manually trimmed to isolate the target speech tasks prior to analysis. Acoustic features were extracted using the open-source openSMILE toolkit (Version 2.5.1) with the eGeMAPS [24]. This standardized configuration provides 88 acoustic features per sample, designed to capture central aspects of speech production, including frequency-, energy/amplitude-, and spectral-related characteristics.

The 88 eGeMAPS features are derived from 18 low-level descriptors (LLDs), which encompass frequency measures (pitch, jitter, formants), energy/amplitude parameters (shimmer, loudness, harmonics-to-noise ratio [HNR]), and spectral descriptors (MFCCs, spectral flux, alpha ratio, Hammarberg index). These LLDs are summarized using functionals such as arithmetic mean and coefficient of variation, generating 36 initial parameters. Additional functionals (e.g., percentiles, mean and standard deviation of slopes) applied to pitch and loudness yield 20 further features. The extended set incorporates seven additional LLDs, primarily cepstral and dynamic parameters, alongside their corresponding functionals and aggregated statistics for voiced and unvoiced regions. Together, these yield 88 features per utterance. The eGeMAPS framework represents an expanded version of the original GeMAPS parameter set, which is widely adopted for standardized voice analysis and has been successfully applied to quantify subtle alterations in speech associated with neurodegenerative and psychiatric disorders [24, 26–28].

These standardized acoustic features served as input for supervised machine learning to determine whether acoustic speech patterns could reliably differentiate cognitively intact individuals by APOE-ε4 genotype (ε4⁺ vs. ε4⁻). The classifier was trained on three data subsets: task-specific speech samples (44 recordings for each task), spontaneous speech data (128 recordings), and a combined dataset comprising all speech types (340 recordings), enabling comparison of model performance across contexts of speech production.

### 2.5 Classifier selection and configuration

Given the characteristics of our dataset, ranging from small (task-specific data) to moderately sized (combined task data), and the high-dimensional feature space with considerable inter-feature correlation, we employed a Random Forest classifier. This ensemble-based method inherently manages high-dimensional data by selecting the most informative features at each split, which helps to reduce the influence of correlated variables. We also used bootstrapping to enhance diversity among trees and reduce variance. The number of estimators was adjusted relative to dataset size to achieve a balance between computational efficiency and variance reduction. Additionally, the maximum tree depth was optimized to prevent overfitting, as excessively deep trees tend to memorize noise and outliers rather than capture generalizable patterns.

### 2.6 Feature preprocessing and evaluation

To reduce noise and redundancy in the input space, we applied a series of preprocessing steps to the extracted features prior to classifier training. First, features exhibiting zero mutual information with the target label were removed, as they offer no predictive value. Next, for each pair of features with a Pearson correlation coefficient greater than 0.9, we retained only the feature with the higher mutual information score to minimize multicollinearity while preserving informative content. These preprocessing steps result in a cleaner, more informative feature set, enabling the classifiers to learn more robust mappings between input features and output labels. To avoid information leakage, all preprocessing operations were performed exclusively on the training data within each split.

Classifier performance was evaluated using stratified random splits of the dataset, with 80% allocated for training and 20% for testing. This procedure was repeated 50 times, and the average performance across all iterations was reported to ensure robustness and mitigate variance due to data partitioning. Results from this baseline setup are presented in section 3.

### 2.7 Feature selection with genetic algorithm (GA)

To enhance classification performance, a GA was employed to select an informative subset of features from the full eGeMAPS. In this framework, each individual in the GA population represents a candidate feature subset, encoded as a binary chromosome, where each bit indicates the inclusion or exclusion of a specific feature. The GA evolves the population through selection, crossover, and mutation, aiming to optimize classification performance.

Key algorithm parameters, including population size and number of generations, were scaled relative to dataset size. For smaller datasets, we employed smaller populations and fewer generations to reduce the risk of overfitting. An early stopping criterion was also implemented: the algorithm terminated if no improvement in performance was observed over a number of generations equal to 20% of the total, thereby avoiding unnecessary computation and overfitting to noise or dataset-specific idiosyncrasies. The crossover probability was set to 0.8, promoting effective exploration of diverse feature subsets. The mutation probability was adaptively adjusted based on dataset size to maintain a balance between exploration of the search space and convergence.

Each individual (i.e., feature subset) was evaluated using a fitness function identical to that used in the baseline classifier setup. Classifier performance was evaluated using the F1 score, which balances precision and sensitivity (recall) by serving as their harmonic mean. This metric was chosen because it penalizes imbalances between precision and recall, offering a more informative and reliable assessment of classification performance than accuracy alone; particularly in scenarios where both false positives and false negatives are consequential. To ensure retention of the best solution, elitism was implemented by tracking the globally best-performing individual throughout the evolutionary process. Upon termination, the feature subset associated with the highest fitness score was retained as the final output.

## Results

### 3.1 Classification Performance Across Tasks

#### 3.1.1 Task Specific

Classifier performance was first evaluated separately for each speech task, with each comprising approximately 44 recordings. To maintain sample size consistency and analytic rigor, a single spontaneous speech task (i.e., participants describing their weekend) was used to represent spontaneous speech. This approach ensured that all included tasks had comparable numbers of recordings.

Figure 1 presents the classification accuracy for each task under two configurations: the baseline model using the full eGeMAPS feature set and the model using only the features selected by the GA. The results indicate that GA-based feature selection improved performance relative to the baseline across all tasks. However, the substantial uncertainties in performance preclude definitive conclusions about which individual task yields superior results. These uncertainties stem from the relatively small size of each dataset, which limits the evaluation sets in the train/test splits and leads to high variance in performance across iterations. This sensitivity to data partitioning underscores the challenges of working with small sample sizes and highlights the importance of reporting uncertainty alongside performance metrics.

**Figure 1.**
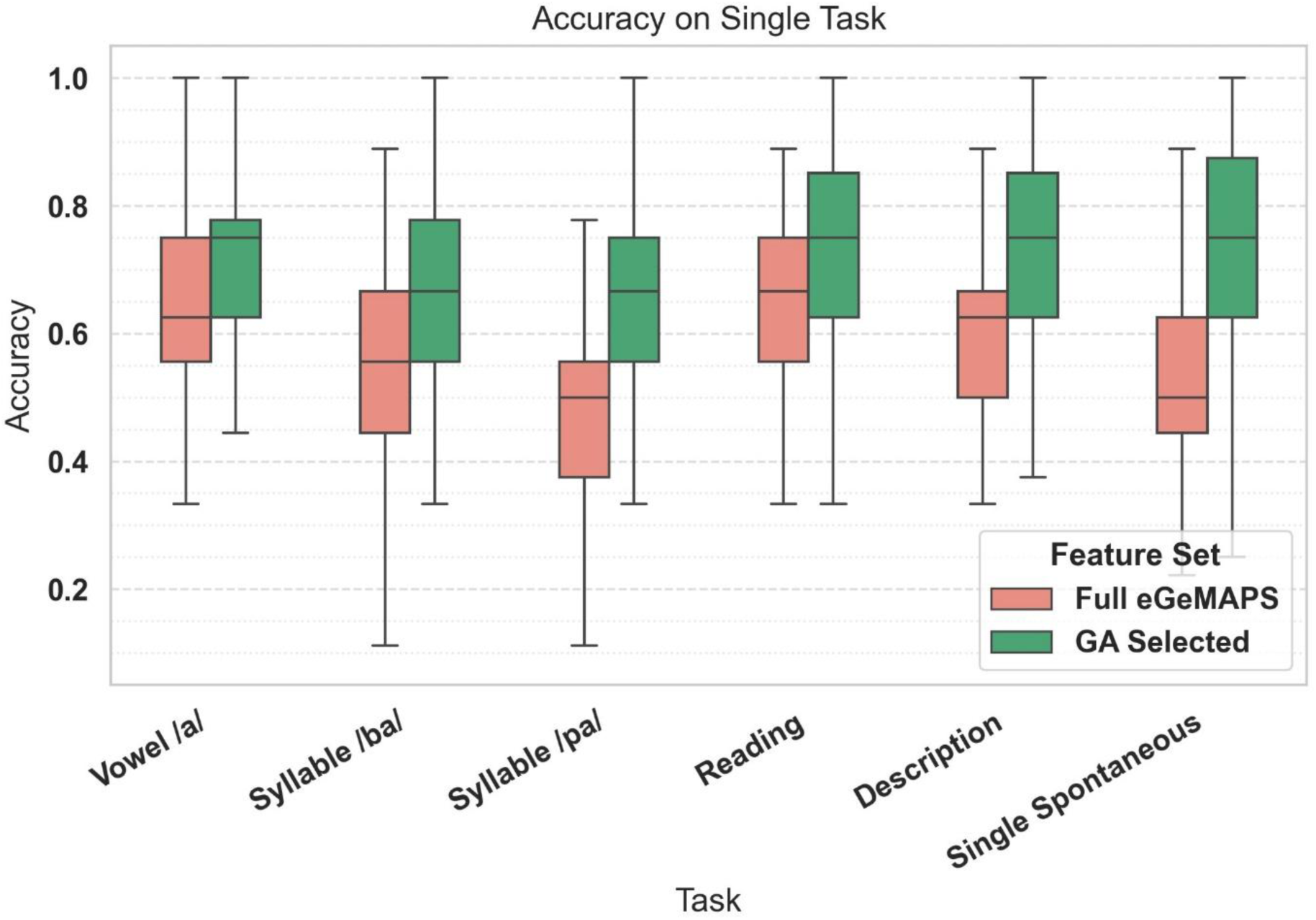
Accuracy of the Random Forest classifier trained on single-task data using either the full eGeMAPS feature set or GA-selected feature set.

#### 3.1.2 Spontaneous Speech Tasks

We further evaluated classifier performance using the combined spontaneous speech dataset, which includes responses from three distinct one-minute tasks in which participants spoke about their weekend, hobbies, and places they have visited. This aggregation yielded a total of 128 recordings. The top panel of Figure 2 illustrates the performance of the classifier across several evaluation metrics: accuracy, precision, sensitivity, specificity, F₁ score and the receiver operating characteristic area under the curve (ROC-AUC), for both the full eGeMAPS feature set (baseline) and the feature subset selected by the GA. A notable improvement in accuracy was observed, with increases of at least 20 percent compared to the accuracies on single task showed in Figure 1, regardless of the feature set used. The larger and more diverse dataset also led to a substantial reduction in performance variability across iterations, enhancing the reliability of the results.

**Figure 2.**
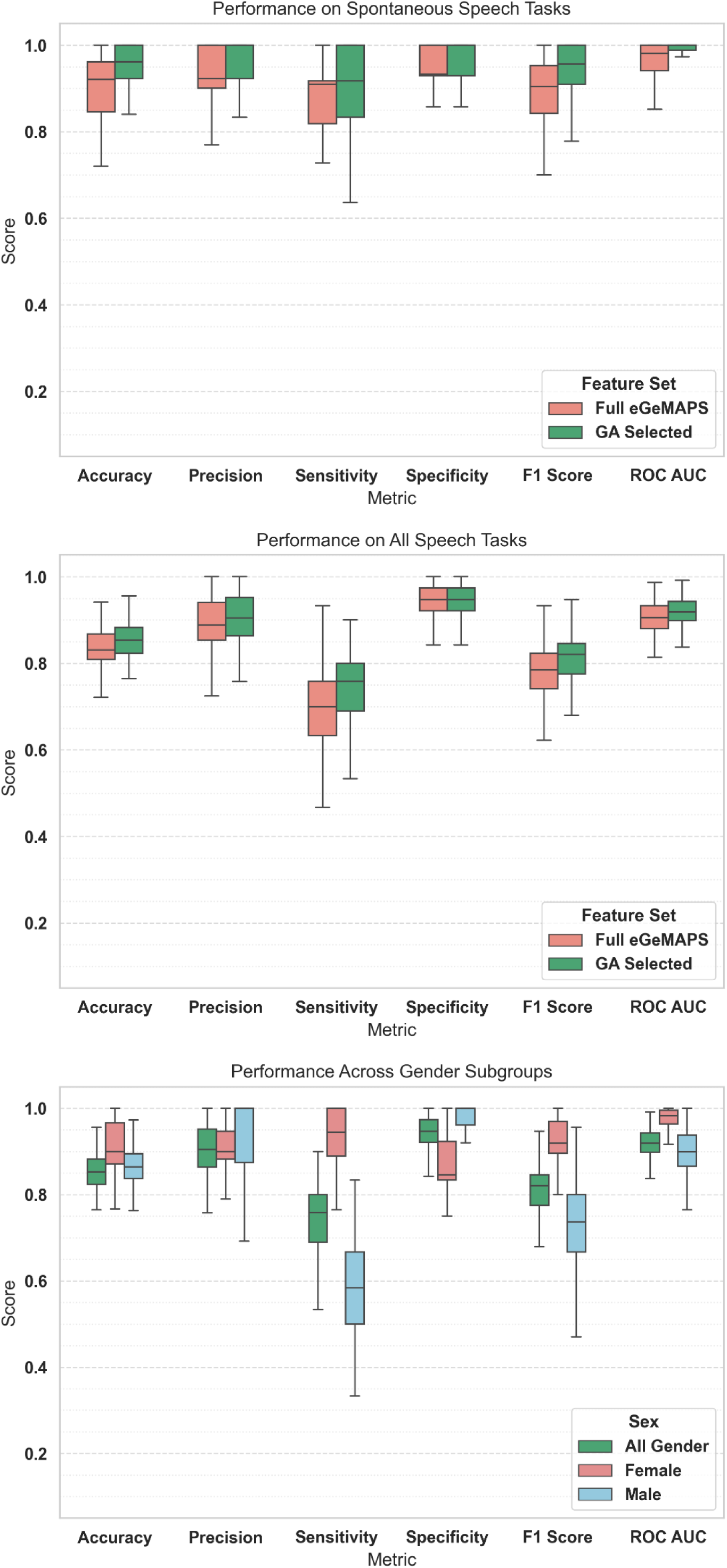
Performance of the Random Forest classifier across task types and gender subgroups. Top panel: spontaneous speech tasks using either the full eGeMAPS feature set or GA-selected feature set. Middle panel: all speech tasks with the same feature sets. Bottom panel: classification performance across gender subgroups on all speech tasks using GA-selected feature sets.

When using GA-selected features, the classification achieved over 90% on all evaluated metrics, underscoring the robustness and discriminative power of the selected features in identifying APOE-ε4 carrier status from spontaneous speech. These findings suggest that spontaneous speech may encode particularly informative acoustic markers that help differentiate between ε4⁺ and ε4⁻ individuals.

#### 3.1.3 Combined tasks

We then assessed the performance of the classifier using the full dataset, which includes speech data from all tasks and totals 340 samples. Training on this aggregated dataset led to a decline in overall performance compared to training exclusively on spontaneous speech data, as shown in the middle panel of Figure 2. The F₁ score of classification with GA-selected features dropped from 0.94 to 0.82. Additionally, the variance of F₁ scores across iterations decreased, indicating improved robustness and generalizability when trained on a larger and more diverse dataset compared to models trained on individual tasks or spontaneous speech alone.

### 3.2 Sex Effect

To investigate potential demographic influences, sex was initially introduced as a categorical feature alongside eGeMAPS features; however, its inclusion did not noticeably enhance classifier performance. Separate classification analyses for male and female subgroups using the combined speech dataset revealed that the classifier performed substantially better among female participants. The bottom panel of Figure 2 compares the classifier performance when trained separately on female-only and male-only data from all tasks using features selected by the GA. The F₁ score improved from 0.81 to 0.93 for females. These results indicate that speech-derived acoustic features are markedly more informative for classifying APOE-ε4 status in females, suggesting possible sex-related differences in the expression of genetic risk for AD at the vocal level.

### 3.3 Selected Features

A diverse spectrum of features including F0, formant frequencies and bandwidths, spectral indices, MFCCs, loudness, voice quality, segment duration, and intensity is selected by a GA paired with a Random Forest classifier across task types (spontaneous and all-tasks) and gender (female, male, all) subgroups as illustrated in the top panel of Figure 3.

**Figure 3.**
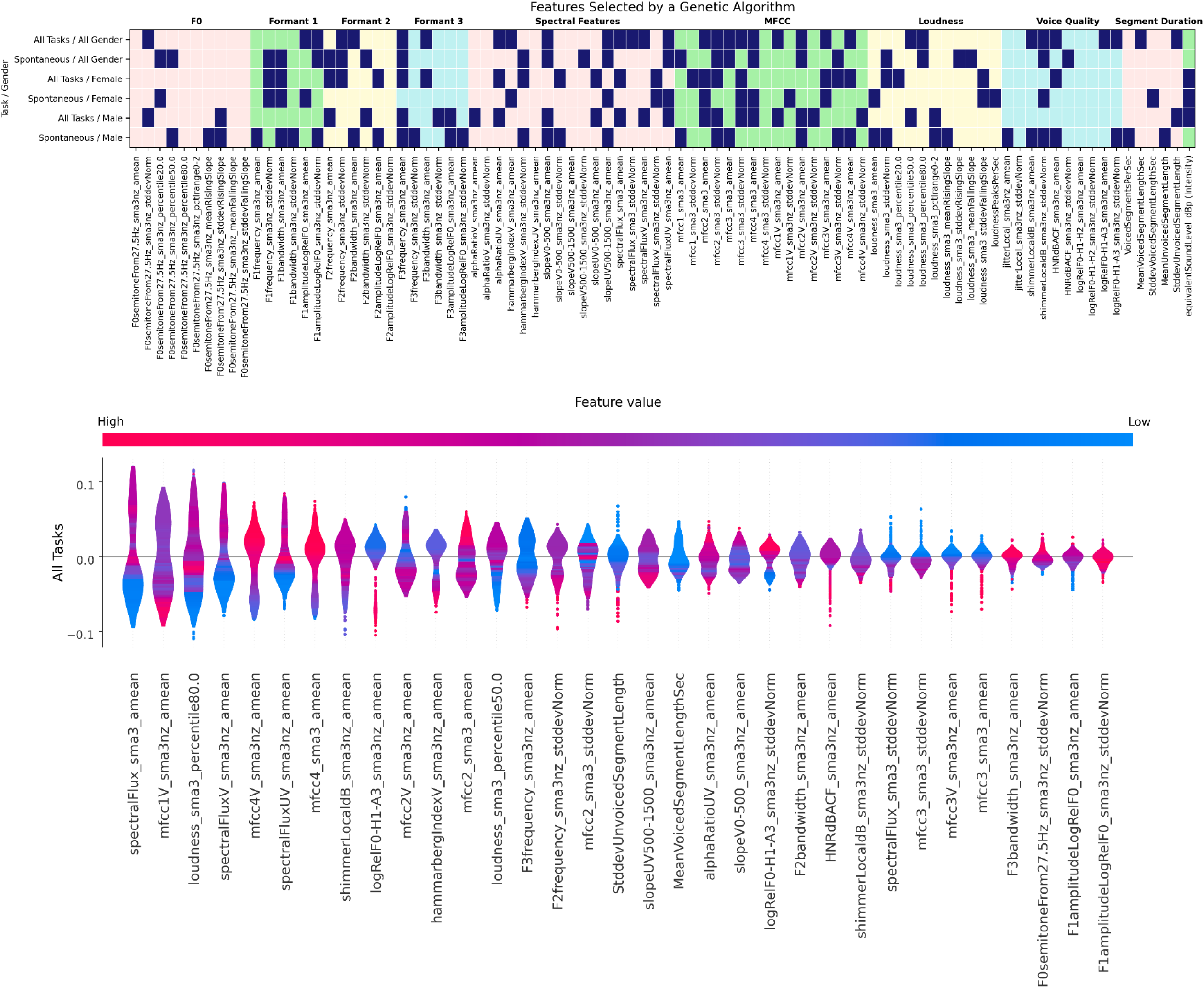
Top panel: features selected by a GA paired with a Random Forest classifier across different task/gender data subsets are displayed by dark blue cells. Bottom panel: SHAP analysis of GA-selected features with a Random Forest classifier trained on all-participant, all-task data. Higher SHAP values indicate a greater predicted probability of APOE ε4 positivity (higher AD risk). Colored areas represent the feature value: blue for lower values and red for higher values. From left to right, the features are ordered by decreasing SHAP importance.

MFCCs show the greatest consistency, universally contributing to APOE-ε4 classification across contexts. F3 features are especially salient for male speakers, with frequent feature selection across both spontaneous and pooled speech tasks, while the intensity feature is more consistently selected in female speakers, highlighting sex-dependent and context-specific risk patterns. Notably, gender– and task-specific trends emerge: F1 frequency and jitter features distinguish male spontaneous speech, while voiced spectral flux variability is prominent in the female subgroup.

To assess how specific acoustic features contribute to classifying APOE-ε4 carriers, we perform SHAP analysis across task types and gender subgroups. Figure 3 (bottom panel) presents the results for the model trained on pooled data. Positive SHAP values indicate an increased probability of ε4+ classification. Low spectral fluxes (spectralFlux(V,UV)_sma3_amean) are predominantly associated with negative SHAP values, indicating a reduced probability of ε4+ classification. In contrast, higher values of MFCC4 (mfcc4(V)_sma3_amean) consistently align with positive SHAP values, strongly increasing the likelihood of ε4+. Additionally, lower loudness values are linked to a decreased probability of ε4+ classification.

## Discussion

This study demonstrates that speech, especially spontaneous speech, serves as a sensitive, noninvasive marker of early APOE-ε4–related vulnerability in cognitively intact older adults. These findings underscore the promise of speech-based digital biomarkers as scalable, ecologically valid, and sensitive to prodromal change for identifying preclinical AD risk and advancing targeted prevention in at-risk populations.

## Feature Selection Patterns Across Task and Gender Subgroups

The integration of feature selection heatmaps and SHAP interpretation strengthens the evidence that the eGeMAPS framework supports a nuanced and comprehensive acoustic characterization of both spontaneous and structured speech in APOE-ε4 research. Application of GAs with Random Forest classifier reliably identified robust markers across multiple domains including MFCCs, formants, temporal indices like articulation rate and segment duration, and spectral properties such as shimmer, and Hammarberg index. These features sensitively capture subtle, preclinical speech alterations associated with APOE-ε4 status, echoing the multidimensional acoustic profiles reported in MCI and AD studies, where combined markers differentiate disease states and monitor progression [29–32].

The selection heatmaps demonstrate that MFCCs are consistently identified as universal markers regardless of task type or gender, supporting their role as core acoustic indicators in both spontaneous and structured speech.

The SHAP analysis provides interpretable insights into how specific acoustic features distinguish APOE-ε4 carriers from non-carriers. MFCC features exhibited differential patterns: while higher mfcc4_sma3_amean values tended to increase the probability of carrier classification, mfcc1v_sma3nz_amean more often reduced it. These divergent trends highlight the context-dependent role of cepstral features and may reflect subtle articulatory or resonance adaptations in genetically at-risk individuals.

Although voice quality metrics appear less often in the algorithmic selection compared to MFCCs or formants, their diagnostic value remains substantiated in the SHAP analysis. Notably, higher values of HNR correlate with more stable glottal function in non-carriers, while elevated shimmer is linked to glottal or laryngeal instability, reflecting established markers observed in clinical cohorts. This finding aligns with previous reports of elevated shimmer in individuals with AD and MCI [31, 33].

Carriers exhibited higher values for intensity, potentially indicating compensatory increases in vocal intensity, effort, or respiratory drive. In contrast, classification of non-carrier status was supported by features such as elevated logRelF0-H1-A3_sma3nz_amean, a sensitive index of glottal quality, along with greater harmonic richness and reduced loudness. These patterns suggest more stable vocal fold function among non-carriers. Additionally, low spectral fluxes (spectralFlux(V,UV)_sma3_amean) strongly decrease ε4+ probability across all task types and gender subgroups.

These findings confirm that risk-related speech alterations are subtle and distributed across multiple acoustic domains, underscoring the need for multivariate feature sets to achieve effective classification. The integration of algorithmic feature selection with SHAP-based interpretability enhances both performance and mechanistic insight, revealing how nuanced acoustic and phonatory adaptations differentiate at-risk individuals from non-carriers even in the absence of overt cognitive symptoms.

## Spontaneous Speech as the Superior Detection Modality

Spontaneous speech tasks yielded a broader and more diverse range of informative acoustic features for distinguishing APOE-ε4 carriers from non-carriers, as evidenced by their prominent selection across gender groups in the feature selection heatmaps. The consistent identification of robust features from spontaneous speech suggests that these tasks may offer particular utility for APOE-ε4 risk classification compared to structured speech tasks such as passage reading and DDK repetitions. This advantage likely stems from the multidimensional cognitive-motor demands of spontaneous speech, which requires the integration of real-time lexical retrieval, working memory, executive control, and articulatory planning. These combined demands may make spontaneous speech uniquely sensitive to subtle preclinical disruptions. Supporting this view, recent systematic studies demonstrate that spontaneous narrative tasks achieve 10–15% higher accuracy than scripted protocols for detecting cognitive impairment [34–36]. Moreover, spontaneous speech offers high ecological validity, as it closely mirrors natural communication contexts and captures fine-grained deficits that remain masked in more constrained, artificial testing conditions.

## Task Heterogeneity and Strategic Feature Selection

Pooling data across all speech tasks did not enhance classification performance and, in some models, led to reduced accuracy compared to task-specific approaches. The feature selection heatmaps demonstrate that different speech tasks yield distinct sets of informative acoustic features, likely due to their varied cognitive and motor demands. As a result, indiscriminate pooling introduces additional heterogeneity, which may obscure salient patterns relevant to APOE-ε4 risk classification. These findings highlight the importance of strategically selecting and optimizing speech tasks, as targeted approaches appear to support early risk detection more effectively than comprehensive, “all-in” pooling strategies. This task-specific advantage is further supported by prior research showing that heterogeneity in speech task demands can dilute the sensitivity of acoustic biomarker models.

Importantly, multivariate combinations of acoustic features consistently provided superior classification performance (higher F_1_ scores) compared to any single marker alone. This trend is also evident in the SHAP summary plot, which demonstrates substantial contributions from multiple, distributed features across MFCCs, spectral, and prosodic domains. These data reinforce the necessity of data-driven, machine learning-based approaches for capturing the subtle, multidimensional patterns characteristic of early disease risk. The reliance on feature combinations highlights that early neuromotor disruption is reflected across a spectrum of acoustic dimensions, rather than isolated to one measurable domain. This principle has been robustly demonstrated in AD and MCI classification studies employing eGeMAPS and related frameworks [37–39]. Notably, the identified acoustic signatures remained stable across both spontaneous and structured tasks, further validating their diagnostic robustness and generalizability. Applying GA-based feature selection effectively addressed the challenges of high-dimensional speech data and yielded substantial performance improvements. The approach consistently prioritized robust prosodic features (such as pitch variability and intonation) and temporal measures (including speech rate and pause duration), markers which have long been recognized as sensitive to neuromotor decline in neurodegenerative disease. These features retained strong predictive value across diverse tasks, supporting their potential as task-invariant, universal biomarkers of APOE-ε4–associated risk.

## Comparison with Prior Studies and Relevance for Early Detection

Recent research applying the GeMAPS/eigenmaps framework to automated speech analysis has consistently demonstrated high diagnostic accuracy for distinguishing MCI and AD [29, 40–42]. For example, García-Gutiérrez et al. [29] reported that eGeMAPS features from spontaneous speech predicted amyloid positivity in MCI with up to 75% accuracy (AUC = 0.79), outperforming conventional neuropsychological tests. Large-scale studies, including the 2024 PROCESS challenge [42], highlight the robustness and scalability of multimodal models incorporating eGeMAPS features, routinely achieving Macro-F1 = 0.5774 for three-group classification of healthy controls, MCI, and Dementia states [42]. Systematic reviews further support the generalizability and real-world feasibility of speech-based biomarkers, demonstrating AUCs of 0.80–0.88 for detecting early AD and MCI across diverse cohorts and in clinical trial enrichment [38, 43, 44].

Building on this foundation, our study extends these findings to an even earlier stage, identifying distinct speech acoustic signatures of AD risk in cognitively normal, APOE-ε4 carriers before any clinical symptom onset. Using eGeMAPS features and advanced machine learning, we achieved classification performance (F1 scores up to 0.90) that matches or exceeds benchmarks set by established MCI and AD studies. Importantly, these early risk signatures were robust to demographic and task variability, and their interpretability (as shown by SHAP analysis) reveals biologically meaningful motor differences that can be detected in prosodic and spectral patterns. These findings demonstrate that multidimensional speech markers are not only effective in identifying symptomatic disease but are also detectable in genetically at risk yet cognitively intact individuals, establishing speech as a powerful preclinical digital biomarker for AD.

## Sex Differences in APOE-ε4 Expression

A striking and consistent finding of this study was the superior classification accuracy for female participants. Across feature sets, and speech tasks, classifiers trained exclusively on female data achieved notably higher F1 scores than those trained on male-only groups. This robust sex difference underscores that biological sex is a key modifier of APOE-ε4–related risk profiles and the expression of disease-relevant speech biomarkers.

Epidemiological studies consistently show that APOE-ε4 confers a greater lifetime risk and earlier onset of AD in women compared to men [45]. Neuroimaging results further support this differential vulnerability, with women exhibiting greater hippocampal volume loss, more rapid cortical thinning, and pronounced reductions in cerebral glucose metabolism [46–48]. In the language domain, female ε4 carriers also appear more susceptible to early verbal memory decline and accelerated language deterioration [49]. This increased clinical vulnerability may manifest as heightened detectability of early neuromotor and cognitive-linguistic changes in the acoustic domain. Prior work by König et al. [50] and others has demonstrated that speech-based models often exhibit higher classification accuracy in female samples, suggesting potential sex-specific acoustic signals associated with preclinical AD. Biological and anatomical factors likely contribute to this effect. Women have shorter vocal tracts and higher fundamental frequencies, making subtle neuromuscular changes more acoustically detectable (see ref. [51] for muscle composition and coordination differences). Our recent research on EMG-derived speech motor control further demonstrates heightened neuromotor change in female ε4 carriers, adding mechanistic depth to behavioral findings [11].

These converging lines of evidence highlight the importance of sex-stratified analyses in digital biomarker development. Pooling male and female data may obscure meaningful sex effects and reduce model sensitivity. The present results support a precision medicine approach in which sex is considered a fundamental biological variable when developing and deploying speech-based screening tools. Tailoring acoustic models to sex-specific vocal and neuromuscular profiles could greatly enhance early AD detection, especially for genetically and clinically high-risk groups such as female APOE-ε4 carriers and inform targeted prevention strategies.

## Limitations and Future Directions

While this study supports speech-derived acoustic features as early, non-invasive biomarkers of Alzheimer’s risk in APOE-ε4 carriers, several limitations warrant consideration. First, the modest sample size constrains generalizability and statistical power. Larger, demographically balanced cohorts are needed to validate findings across diverse populations and better model the influence of variables like age, sex, ethnicity, and education. Second, this analysis did not examine interactions between APOE status and other risk factors (e.g., vascular health, and other comorbidities), which may independently or interactively shape speech phenotypes and neurodegeneration. Including these variables in future models will help disentangle complex real-world influences and reduce confounding effects. Third, the cross-sectional design limits causal inference. Longitudinal studies are essential to assess whether the identified speech signatures precede cognitive decline or track disease progression. Future research should also integrate multimodal data by combining speech with other digital, physiological, or imaging biomarkers to reveal convergent early disease signatures and clarify underlying neural mechanisms. Finally, for clinical translation, speech-based models must be validated in large, independent, and naturalistic cohorts including remote and passive monitoring via smartphones and telehealth.

## Conclusion

This study demonstrates that spontaneous speech, with its cognitive-motor complexity, is highly sensitive for detecting early neurobiological changes associated with APOE-ε4. Machine learning–based feature selection proved vital for extracting discriminative signals from rich acoustic data. Using the eGeMAPS framework and robust modeling, we show that acoustic features, especially from spontaneous speech, can non-invasively identify APOE-ε4 status in cognitively normal older adults. These early vocal signatures reflect neuromuscular and cognitive vulnerability and highlight speech analysis as a scalable, behaviorally grounded digital biomarker for preclinical AD. Our findings underscore the value of incorporating motor-based speech assessments into future risk prediction frameworks for at-risk populations.

## Data Availability

Because audio recordings of participants' speech may be considered potentially identifiable and could compromise confidentiality, we do not share raw audio files. Instead, we provide Excel files containing the processed output used in the data analyses. All data generated in the present study are available from the authors upon reasonable request.

## Acknowledgements

We gratefully acknowledge all participants who generously contributed their time and effort to this study. Their willingness to share their experiences and complete the speech tasks made this research possible. We further extend our gratitude to the families and caregivers who supported participant involvement, and to Hannah Corinne Weibley and Vijayaraghavan Gayathri for their invaluable contributions to participant enrollment and clinical data collection.

## Author Contribution

M.T. contributed to data analysis and manuscript writing. M.D. reviewed the manuscript and provided feedback on data analysis and statistical methodology. J.R.G. reviewed the manuscript and provided feedback. K.P.C. contributed to perceptual evaluation of speech samples, reviewed the manuscript, and provided feedback. B.D.R. supported data collection, contributed to IRB submission and approval, reviewed the manuscript, and provided feedback. N.V.B. assisted with data collection, reviewed the manuscript, and provided feedback. M.Tk. assisted with post-processing of audio signals for acoustic analysis. D.H.S. contributed to participant characterization, manuscript review, and feedback. S.E.A. was involved in participant characterization and reviewed and provided feedback on the manuscript. M.E. served as the Principal Investigator and played a leading role in study conceptualization and design, data collection, data analysis, perceptual evaluation of speech samples, interpretation of findings, and manuscript writing. All authors contributed to the article and approved the submitted version

## Consent Statement

Recruitment and consent procedures adhered to Mass General Brigham Healthcare (MGH) and HIPAA regulations. The study was approved by the MGB Institutional Review Board (IRB Protocol #2021P001460).

Before participating in testing, administrators thoroughly explained the experimental tasks and procedures, addressing any questions participants had. Relevant consent forms were provided, and each form was reviewed point by point with potential participants by a research staff member prior to obtaining their signature. Participants were explicitly informed of their right to withdraw from the study at any time during the experiment without any consequences and all participants provided written informed consent prior to participation.

## Appendix 1

*Sustained phonation task:* Participants were asked to take a deep breath and prolong the vowel /a/ for as long as possible, at their typical pitch and loudness. The sustained phonation task is widely used in voice assessment and therapy, as it provides critical insights into glottic function (i.e., vocal fold closure) and respiratory-phonatory coordination [52], while offering a controlled context for precise acoustic measurements. This task is particularly valuable for acoustic analysis because it yields a stable and continuous vocal signal, minimizing variability from pitch modulation, loudness changes, and articulatory influences seen in connected speech. By reducing the effects of coarticulation and prosodic variation, sustained phonation enables precise extraction of key acoustic features such as fundamental frequency (F0), jitter, and shimmer. These acoustic parameters commonly used to assess voice quality and detect vocal pathology. In addition to its diagnostic utility, the simplicity of the task facilitates standardized comparisons across individuals and time points.

*Syllable repetitions in an oral diadochokinetic paradigm:* Oral DDK tasks are clinical tools used to assess oromotor coordination, speed, and precision by measuring rapid syllable repetition. These tasks include Alternating Motion Rate (AMR) tasks which involve the rapid and accurate repetition of a single syllable on a single breath and are commonly used to evaluate execution-level motor speech control. In this study, we will use the syllables /ba/ and /pa/, which differ in voicing (i.e., /ba/ being voiced and /pa/ voiceless) while both require bilabial closure. These differences allow us to examine subtle variations in articulatory timing and control. Impaired AMR performance, such as slowed rate or increased variability, can indicate neuromuscular dysfunction, commonly observed in disorders like dysarthria. AMR tasks are also sensitive to subtle changes in articulatory speed and stability across age groups or neurological conditions [53, 54]. While these tasks do not fully capture the complexity of spontaneous speech, they provide a controlled and quantifiable measure of speech motor performance, offering valuable insights into oromotor integrity and efficiency.

*Passage reading:* In this study participants were asked to read the “Bamboo Passage” which is a 60-word paragraph containing voiced consonants (e.g., /b/ and /d/) at word and phrase boundaries to eliminate misidentification of pauses that can occur during voiceless stops. This task provides sensitive measures of motor speech fluency including speaking rate, pause duration, and articulatory precision and has been used in dysarthria research to identify errors in fine-motor speech sounds, with multiple studies showing it can distinguish individuals with early-stage ALS from healthy controls [55, 56]. The passage’s standardized content enables semi-automated, reliable tracking of subtle speech changes for diagnosis, and disease progression and treatment response in clinical trials. In addition to capturing motor deficits, performance on the task reflects cognitive functions such as sustained attention, visual discrimination, processing speed, and linguistic processing (phonology, semantics, syntax). While repeated use may introduce practice effects and its structure may not fully represent spontaneous speech, the Bamboo Passage remains a robust tool for identifying early motor speech changes and developing speech-based biomarkers especially when used alongside naturalistic tasks for a comprehensive assessment.

*Picture description task:* Picture description relies less on episodic memory and involves cognitive processes including visual attention, sustained attention, object recognition, lexical retrieval, phonological encoding, semantic and syntactic processes, and self-monitoring.

The Western Aphasia Battery (WAB) Picnic Scene is a standardized picture description task widely used in clinical and research settings to elicit connected speech for assessing speech motor fluency and language abilities in individuals with neurological conditions such as aphasia, ALS, and dementia. By prompting participants to describe a complex visual scene involving multiple characters and activities (e.g., kite flying, picnicking, sailing), the task relies less on episodic memory but systematically engages visual attention, object recognition, lexical retrieval, phonological encoding, syntactic structuring, and semantic integration. The task’s fixed content supports consistent cross-group comparisons and is well-suited for automated speech analysis, enabling objective measurement of acoustic features and error patterns to detect early markers of neurological decline.

*Spontaneous speech samples:* Participants were asked to speak for one minute on each of the following prompts: (1) describe what a typical weekend looks like for you, (2) describe your hobbies and activities, and (3) describe a place or country you have visited and particularly enjoyed. These open-ended tasks are designed to tap into natural language production abilities, engaging a broad array of cognitive-linguistic and speech motor processes including lexical retrieval, syntactic construction, articulation, and prosodic modulation. Unlike structured tasks, spontaneous speech offers a more ecologically valid window into real-world communication, making it especially valuable for detecting subtle deficits that may not emerge in constrained contexts. When combined with automated speech analysis, these samples can be systematically evaluated for acoustic and linguistic features, enhancing sensitivity to early cognitive or neuromotor changes, supporting reproducible and scalable analysis, and enabling longitudinal tracking of disease progression or treatment response in clinical populations.

## References

[1] Sperling RA, Aisen PS, Beckett LA, et al (2011) Toward defining the preclinical stages of Alzheimer’s disease: Recommendations from the National Institute on Aging-Alzheimer’s Association workgroups on diagnostic guidelines for Alzheimer’s disease. Alzheimer’s Dement 7:280–292. 10.1016/j.jalz.2011.03.003

[2] Raber J, Huang Y, Ashford JW (2004) ApoE genotype accounts for the vast majority of AD risk and AD pathology. Neurobiol Aging 25:641–650. 10.1016/j.neurobiolaging.2003.12.023

[3] Feskens EJM, Havekes LM, Kalmijn S, et al (1994) Apolipoprotein e4 allele and cognitive decline in elderly men. BMJ 309:1202–1206. 10.1136/bmj.309.6963.1202

[4] Buchman AS, Boyle PA, Wilson RS, et al (2009) Apolipoprotein E e4 Allele is Associated With More Rapid Motor Decline in Older Persons. Alzheimer Dis Assoc Disord 23:63–69. 10.1097/WAD.0b013e31818877b5

[5] Henderson AS, Jorm AF, Korten AE, et al (1995) Apolipoprotein E allele∈ 4, dementia, and cognitive decline in a population sample. Lancet 346:1387–1390. 10.1016/S0140-6736(95)92405-1

[6] Dik MG, Jonker C, Bouter LM, et al (2000) APOE-ε4 is associated with memory decline in cognitively impaired elderly. Neurology 54:1492–1497. 10.1212/WNL.54.7.1492

[7] El Haj M, Antoine P, Amouyel P, et al (2016) Apolipoprotein E (APOE) ε4 and episodic memory decline in Alzheimer’s disease: A review. Ageing Res Rev 27:15–22. 10.1016/j.arr.2016.02.002

[8] Sun J, Zhu Z, Chen K, et al (2020) APOE ε4 allele accelerates age-related multi-cognitive decline and white matter damage in non-demented elderly. Aging (Albany NY) 12:12019–12031. 10.18632/aging.103367

[9] Reas ET, Laughlin GA, Bergstrom J, et al (2019) Effects of APOE on cognitive aging in community-dwelling older adults. Neuropsychology 33:406–416. 10.1037/neu0000501

[10] Li W, Qiu Q, Sun L, et al (2019) Short-term adverse effects of the apolipoprotein E ε4 allele over language function and executive function in healthy older adults. Neuropsychiatr Dis Treat 15:1855–1861. 10.2147/NDT.S183064

[11] Eshghi M, Rong P, Dadgostar M, et al (2025) APOE-ε4 Modulates Facial Neuromuscular Activity in Nondemented Adults: Toward Sensitive Speech-Based Diagnostics for Alzheimer’s Disease. medRxiv. 2025 Apr 29:2025-04.

[12] Liampas I, Demiri S, Siokas V, et al (2025) Apolipoprotein E Alleles and Motor Signs in Older Adults with Alzheimer’s Dementia. Int J Mol Sci 26:8562. 10.3390/ijms26178562

[13] Dadgostar M, Hanford LC, Green JR, et al (2025) Kinematic Correlates of Early Speech Motor Changes in Cognitively Intact APOE-ε4 Carriers: A Preliminary Study Using a Color-Word Interference Task. medRxiv. 2025:2025–06.

[14] Fraser KC, Meltzer JA, Rudzicz F (2015) Linguistic features identify Alzheimer’s disease in narrative speech. J Alzheimer’s Dis 49:407–422. 10.3233/JAD-150520

[15] Szatloczki G, Hoffmann I, Vincze V, et al (2015) Speaking in Alzheimer’s disease, is that an early sign? Importance of changes in language abilities in Alzheimer’s disease. Front Aging Neurosci 7:. 10.3389/fnagi.2015.00195

[16] Filiou R-P, Bier N, Slegers A, et al (2020) Connected speech assessment in the early detection of Alzheimer’s disease and mild cognitive impairment: a scoping review. Aphasiology 34:723–755. 10.1080/02687038.2019.1608502

[17] Ivanova O, Meilán JJG, Martínez-Sánchez F, et al (2022) Discriminating speech traits of Alzheimer’s disease assessed through a corpus of reading task for Spanish language. Comput Speech Lang 73:101341. 10.1016/j.csl.2021.101341

[18] Petti U, Baker S, Korhonen A (2020) A systematic literature review of automatic Alzheimer’s disease detection from speech and language. J Am Med Informatics Assoc 27:1784–1797. 10.1093/jamia/ocaa174

[19] Mueller KD, Hermann B, Mecollari J, Turkstra LS Connected speech and language in mild cognitive impairment and Alzheimer’s disease: A review of picture description tasks. J Clin Exp Neuropsychol 40:917–939. 10.1080/13803395.2018.1446513

[20] Hason L, Krishnan S (2022) Spontaneous speech feature analysis for alzheimer’s disease screening using a random forest classifier. Front Digit Heal 4:901419. 10.3389/fdgth.2022.901419

[21] Martínez-Nicolás I, Llorente TE, Martínez-Sánchez F, Meilán JJG (2021) Ten Years of Research on Automatic Voice and Speech Analysis of People With Alzheimer’s Disease and Mild Cognitive Impairment: A Systematic Review Article. Front Psychol 12:. 10.3389/fpsyg.2021.620251

[22] Skoog I, Hörder H, Frändin K, et al (2016) Association between APOE genotype and change in physical function in a population-based Swedish cohort of older individuals followed over four years. Front Aging Neurosci 8:. 10.3389/fnagi.2016.00225

[23] Hilsabeck RC, Keller JN, Henry ML, et al (2025) Development and classification accuracy of an automated cognitive screening tool combining working memory and connected speech tasks for early detection of cognitive impairment in primary care. Alzheimer’s Dement Transl Res Clin Interv 11:. 10.1002/trc2.70145

[24] Eyben F, Scherer KR, Schuller BW, et al (2016) The Geneva Minimalistic Acoustic Parameter Set (GeMAPS) for Voice Research and Affective Computing. IEEE Trans Affect Comput 7:190–202. 10.1109/TAFFC.2015.2457417

[25] Yorkston KM, Beukelman DR (1981) Communication Efficiency of Dysarthric Speakers as Measured by Sentence Intelligibility and Speaking Rate. J Speech Hear Disord 46:296–301. 10.1044/jshd.4603.296

[26] Gutz SE, Wang J, Yunusova Y, Green JR (2019) Early Identification of Speech Changes Due to Amyotrophic Lateral Sclerosis Using Machine Classification. In: Interspeech 2019. ISCA, pp 604–608

[27] Turnispeed J (2024) Evaluating the Performance of eGeMAPS Features in Depression Detection Using E-Daic Subsets. Northern Illinois University

28. de Boer JN, Voppel AE, Brederoo SG, et al (2023) Acoustic speech markers for schizophrenia-spectrum disorders: a diagnostic and symptom-recognition tool. Psychol Med 53:1302–1312. 10.1017/S0033291721002804

[29] García-Gutiérrez F, Marquié M, Muñoz N, et al (2023) Harnessing acoustic speech parameters to decipher amyloid status in individuals with mild cognitive impairment. Front Neurosci 17:1221401. 10.3389/fnins.2023.1221401

[30] Choi ASG, Kim J, Kim S, et al (2024) Crosslinguistic Acoustic Feature-based Dementia Classification using Advanced Learning Architectures. InProceedings of the Fifth Workshop on Resources and ProcessIng of linguistic, para-linguistic and extra-linguistic Data from people with various forms of cognitive/psychiatric/developmental impairments@ LREC-COLING 2024. ELRA and ICCL, Torino, Italia, pp 95–100

[31] Parlak MM, Saylam G, Babademez MA, et al (2023) Voice analysis results in individuals with Alzheimer’s disease: How do age and cognitive status affect voice parameters?. Brain Behav 13:e3271. 10.1002/brb3.3271

[32] Qi X, Zhou Q, Dong J, Bao W (2023) Noninvasive automatic detection of Alzheimer’s disease from spontaneous speech: a review. Front Aging Neurosci 15:1224723. 10.3389/fnagi.2023.1224723

[33] Mahon E, Lachman ME (2022) Voice biomarkers as indicators of cognitive changes in middle and later adulthood. Neurobiol Aging 119:22–35. 10.1016/j.neurobiolaging.2022.06.010

[34] Chen J, Ye J, Tang F, Zhou J (2021) Automatic detection of alzheimer’s disease using spontaneous speech only. Interspeech 2021:3830–3834. 10.21437/interspeech.2021-2002

[35] Luz S, Haider F, de la Fuente Garcia S, et al (2021) Editorial: Alzheimer’s dementia recognition through spontaneous speech. Front Comput Sci 3:780169. 10.3389/fcomp.2021.780169

[36] Syed MSS, Syed ZS, Lech M, Pirogova E (2020) Automated Screening for Alzheimer’s Dementia Through Spontaneous Speech. In: Interspeech 2020. ISCA, pp 2222–2226

[37] Saeedi S, Hetjens S, Grimm MOW, Barsties V., Latoszek B (2024) Acoustic Speech Analysis in Alzheimer’s Disease: A Systematic Review and Meta-Analysis. J Prev Alzheimer’s Dis 11:1789– 1797. 10.14283/jpad.2024.132

[38] Qi W, Zhu X, Wang B, et al (2025) Alzheimer’s disease digital biomarkers multidimensional landscape and AI model scoping review. npj Digit Med 8:366. 10.1038/s41746-025-01640-z

[39] Fristed E, Skirrow C, Meszaros M, et al (2022) Leveraging speech and artificial intelligence to screen for early Alzheimer’s disease and amyloid beta positivity. Brain Commun 4:fcac231. 10.1093/braincomms/fcac231

[40] Huang L, Yang H, Che Y, Yang J (2024) Automatic speech analysis for detecting cognitive decline of older adults. Front Public Heal 12:1417966. 10.3389/fpubh.2024.1417966

[41] Bang J, Han S, Kang B (2024) Alzheimer’s disease recognition from spontaneous speech using large language models. ETRI J 46:96–105. 10.4218/etrij.2023-0356

[42] Chi L, Sharma A, Gebhardt A, Colonel JT (2025) Predicting Cognitive Decline: A Multimodal AI Approach to Dementia Screening from Speech. In2025 IEEE International Conference on AI and Data Analytics (ICAD) 2025 Jun 24 (pp. 1–8). IEEE. https://10.1109/ICAD65464.2025.11114053

[43] Alexopoulou Z-S, Köhler S, Mallick E, et al (2025) Speech-based digital cognitive assessments for detection of mild cognitive impairment: Validation against paper-based neurocognitive assessment scores. J Alzheimer’s Dis 13872877251343296. 10.1177/13872877251343296

[44] Oh C, Park MS (2025) Unveiling 30 years of research on speech biomarker of dementia using text mining. Digit Heal 11:20552076251360900. 10.1177/20552076251360901

[45] Altmann A, Tian L, Henderson VW, et al (2014) Sex modifies the APOE-related risk of developing Alzheimer disease. Ann Neurol 75:563–573. 10.1002/ana.24135

[46] Holland D, Desikan RS, Dale AM, McEvoy LK (2013) Higher Rates of Decline for Women and Apolipoprotein E ε4 Carriers. Am J Neuroradiol 34:2287–2293. 10.3174/ajnr.A3601

[47] Buckley RF, Mormino EC, Rabin JS, et al (2019) Sex differences in the association of global amyloid and regional tau deposition measured by positron emission tomography in clinically normal older adults. JAMA Neurol 76:542. 10.1001/jamaneurol.2018.4693

[48] Mosconi L, Berti V, Quinn C, et al (2017) Sex differences in Alzheimer risk: Brain imaging of endocrine vs chronologic aging. Neurology 89:1382–1390. 10.1212/WNL.0000000000004425

[49] Sundermann EE, Biegon A, Rubin LH, et al (2016) Better verbal memory in women than men in MCI despite similar levels of hippocampal atrophy. Neurology 86:1368–1376. 10.1212/WNL.0000000000002570

[50] König A, Linz N, Tröger J, et al (2018) Fully automatic speech-based analysis of the semantic verbal fluency task. Dementia and geriatric cognitive disorders. Dement Geriatr Cogn Disord 45:198–209. 10.1159/000487852

[51] Hunter SK (2009) Sex differences and mechanisms of task-specific muscle fatigue. Exerc Sport Sci Rev 37:113–122. 10.1097/JES.0b013e3181aa63e2

[52] Maslan J, Leng X, Rees C, et al (2011) Maximum phonation time in healthy older adults. J Voice 25:709–713. 10.1016/j.jvoice.2010.10.002

[53] Ben-David BM, Icht M (2017) Oral-diadochokinetic rates for Hebrew-speaking healthy ageing population: Non-word versus real-word repetition. Int J Lang Commun Disord 52:301–310. 10.1111/1460-6984.12272

[54] Tafiadis D, Zarokanellou V, Prentza A, et al (2022) Diadochokinetic rates in healthy young and elderly Greek-speaking adults: The effect of types of stimuli. Int J Lang & Commun Disord 57:1085–1097. 10.1111/1460-6984.12747

[55] Yunusova Y, Green JR, Wang J, et al (2011) A Protocol for Comprehensive Assessment of Bulbar Dysfunction in Amyotrophic Lateral Sclerosis (ALS). J Vis Exp. 10.3791/2422

[56] Simmatis LE, Robin J, Pommée T, et al (2023) Validation of automated pipeline for the assessment of a motor speech disorder in amyotrophic lateral sclerosis (ALS). Digit Heal 9:20552076231219104. 10.1177/20552076231219102

